# Patterns of blunt and cigar use in the United States, 2015-2019

**DOI:** 10.1101/2024.02.26.24303391

**Authors:** Jessica K Jensen, Ollie Ganz, Marisa Tomaino, Allison M Glasser, Kymberle Sterling, Cristine D Delnevo, Michelle T Bover Manderski

## Abstract

**Background:** The use of cigars for blunts (i.e., cannabis rolled in cigar paper) is well-documented; proportions of cigar and blunt use and associated characteristics are less studied.

**Methods:** Pooled data from the 2015-2019 National Survey on Drug Use and Health (NSDUH) were analyzed in 2023. Respondents aged 12+ who reported past 30-day cigar use were categorized into three mutually exclusive use categories: (1) exclusively cigars, (2) exclusively blunts, and (3) both cigars and blunts. We examined associations between cigar-blunt use category and sociodemographic characteristics.

**Results:** Among people 12 and older who reported past 30-day cigar use, 48.6% (95% CI=47.6-49.6) reported exclusive cigar use; 44.3% (95% CI=43.3-45.3) reported exclusive blunt use; and 7.2% (95% CI=6.8-7.6) reported cigars and blunts. The prevalence differed by age, with exclusively blunts most prevalent among youth (72.5% [95% CI=70.7-74.3]) and young adults (62.4% [95% CI=61.4-63.5]), and exclusively cigars most prevalent among adults 26+ (61.2% [95% CI=59.8-62.5]). Exclusive blunt users smoked more days in the past month (17.5; 95% CI=16.8-18.2), compared to 13.8 days (95% CI=13.2-14.4) for cigar and blunt users, and 7.7 days (95% CI=7.5-8.0) for exclusive cigar users. There were significant differences in characteristics, with exclusive blunt use more prevalent among female (41.6%; 95% CI=40.3-42.9) and Hispanic (18.2%; 95% CI=17.3-19.2) participants.

**Conclusions:** Exclusive blunt use was the most prevalent pattern of past-30-day cigar use among youth and young adults. Those who use cigars as blunts smoke more cigars per month, suggesting this may be an important group for additional education and policy efforts.

---

Concurrent cigar and blunt (i.e., replacing some or all of the tobacco within a cigar with cannabis^1^) use is common within the United States (U.S.)^2^, with some data suggesting that blunt use is more popular than cigar use among sub-populations, including Black and male young adults.^3^ Cigar-blunt co-use may occur through the use of separate products at the same time, on the same day, or on different days within the past month.^4^ There is some evidence of a synergistic relationship, in which a greater high is experienced when using both products compared to either product individually. This enhanced effect is often attributed to interactions between tetrahydrocannabinol and nicotine^5,6^ and may contribute to increased dependence of both products.^7^ The act of chasing blunt use with cigars is also common as a way of disguising or enhancing blunt use.^8^

Cigars and cannabis are often perceived as safer products than cigarettes, with blunts perceived as safer than cigars due to the perceived removal of tobacco and nicotine.^9–11^ However, concurrent use is associated with increased nicotine and cannabis dependence,^7,12^ lower intentions to quit,^13^ less successful cessation attempts,^14^ and increased risk for negative health outcomes.^13^ Additionally, exclusive use of either cigars or blunts individually is associated with increased likelihood of future use of the other product.^15,16^

Past studies have noted that older adults and males tend to exclusively use cigars,^17^ while blunt and concurrent cigar-blunt use are more prevalent among young adults and Black youth^18^ and adults.^19^ Other studies examining sociodemographic characteristics associated with blunt use irrespective of cigar use status, found that females and lesbian, gay, or bisexual adults were at increased odds of using blunts compared to their counterparts.^9,18,20^ However, patterns of concurrent and exclusive cigar and blunt use have not been well studied. As such, we sought to identify the (1) proportion of cigar users that consume blunts and (2) sociodemographic characteristics across cigar-blunt use categories. We focus specifically on cigar-blunt co-use as these represent combusted products with higher rates of concurrent use.

## METHODS

### Data Source

We analyzed pooled data from the National Survey of Drug Use and Health (NSDUH) 2015-2019 public-use files (N=282,768 participants aged 12 and older). We further restricted our sample to those who had reported using cigars in the past 30 days or who could be inferred to have used them by their reported blunt use (N=25,004).

### Sample

We categorized respondents into three mutually exclusive groups based on their reported cigar and blunt use in the past 30 days: exclusively cigars, exclusively blunts, and both cigars and blunts. Participants who reported no past 30-day blunt use were categorized as exclusively using cigars. Those who responded that all the cigars they had smoked in the past 30 days contained cannabis were categorized as exclusively using blunts. Some participants (n=10,829) reported blunt use but not cigar use and were also categorized as exclusively using blunts. Those who reported that they smoked some but not all their cigars with cannabis in the past 30 days were categorized as using both cigars and blunts. When conflicting responses to cigar and blunt questions caused a respondent to qualify for more than one group (n=147), such as reporting having never used cannabis and also reporting having smoked a cigar with cannabis in it in the past 30 days, we used NSDUH’s questions about disagreement to determine their categorization (see Supplemental Table 1). When this information was missing or unclear, respondents were excluded from the sample (n=12).

### Measures

#### Frequency of Use

The primary outcome of interest was frequency of cigar use, defined as the number of days used within the past 30 days. Participants who reported past 30-day cigar and/or blunt use were asked to report the number of days they smoked all or part of a cigar and/or blunt. For participants responding inconsistently across the two items, the higher reported number was used.

#### Sociodemographic and substance use characteristics

Sociodemographic characteristics included age, sex, race/ethnicity, sexual identity, education, poverty status, and psychological distress, categorized as shown in Table 1. The cigar brand most used in the past 30 days was assessed among participants reporting past 30-day cigar use and did not include participants who did not report cigar use (i.e., the n=10,829 who only reported blunt use). We reported the five most prevalent brands for each cigar-blunt group separately and combined all other specified brands into the category “other brands in survey.” We kept unspecified brands separate as they were defined in the original data. We used self-reported time since last smoked to create indicators for whether participants had smoked cigarettes and cannabis or hashish in the past 30 days.

**Table 1.**
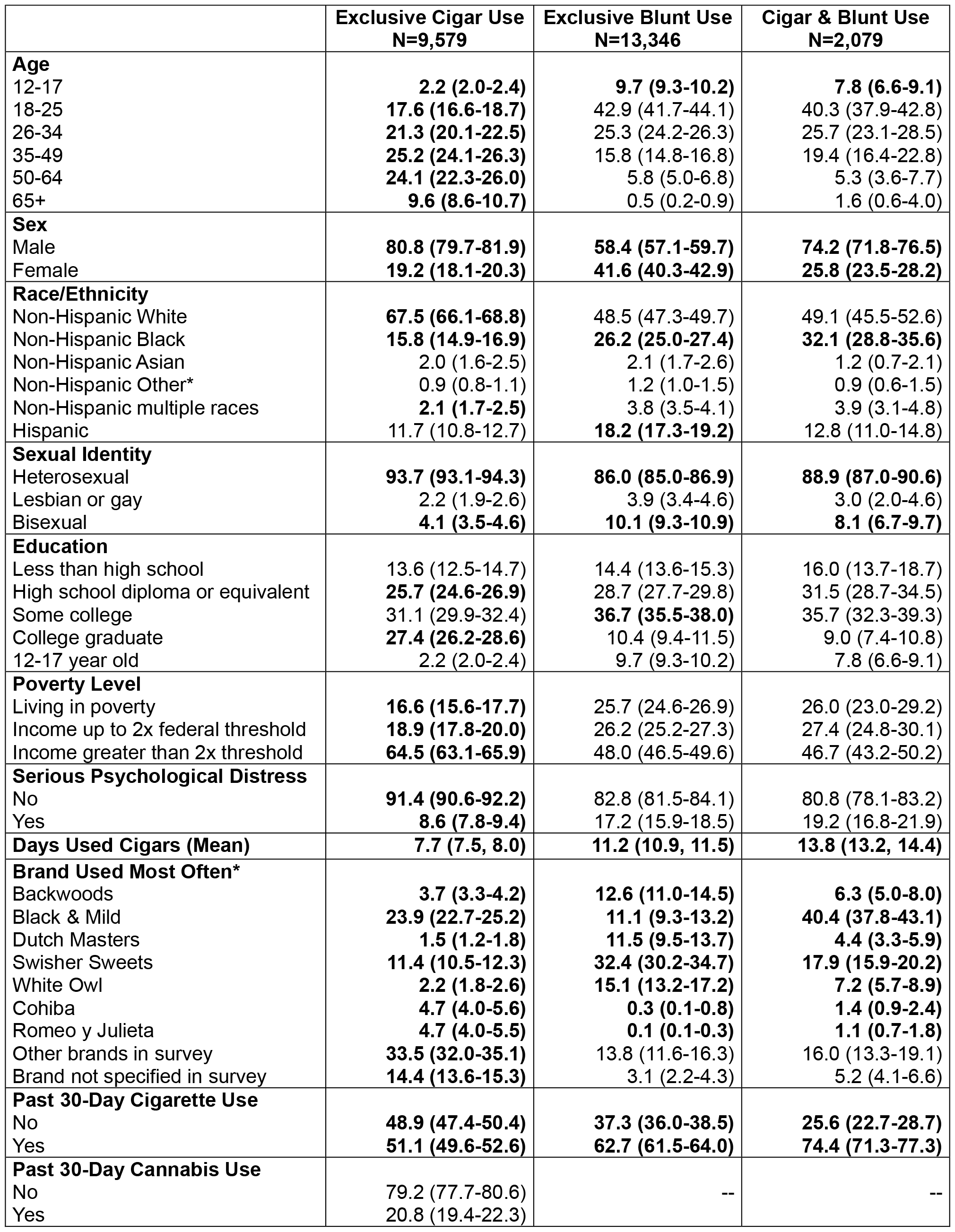
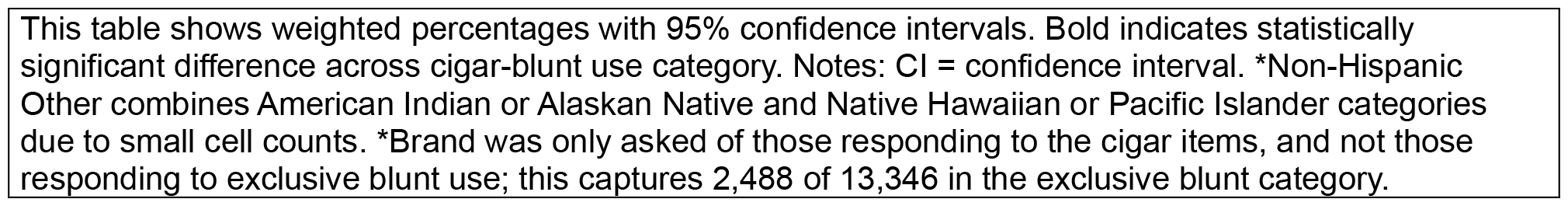
Sample Characteristics by Cigar-Blunt Use Status, % (95% CI)

| **Table 1. Sample Characteristics by Cigar-Blunt Use Status, % (95% CI)** |  |  |  |
| --- | --- | --- | --- |
|  | **Exclusive Cigar Use**<br>**N=9,579** | **Exclusive Blunt Use**<br>**N=13,346** | **Cigar & Blunt Use**<br>**N=2,079** |
| **Age** |  |  |  |
| 12-17 | **2.2 (2.0-2.4)** | **9.7 (9.3-10.2)** | **7.8 (6.6-9.1)** |
| 18-25 | **17.6 (16.6-18.7)** | 42.9 (41.7-44.1) | 40.3 (37.9-42.8) |
| 26-34 | **21.3 (20.1-22.5)** | 25.3 (24.2-26.3) | 25.7 (23.1-28.5) |
| 35-49 | **25.2 (24.1-26.3)** | 15.8 (14.8-16.8) | 19.4 (16.4-22.8) |
| 50-64 | **24.1 (22.3-26.0)** | 5.8 (5.0-6.8) | 5.3 (3.6-7.7) |
| 65+ | **9.6 (8.6-10.7)** | 0.5 (0.2-0.9) | 1.6 (0.6-4.0) |
| **Sex** |  |  |  |
| Male | **80.8 (79.7-81.9)** | **58.4 (57.1-59.7)** | **74.2 (71.8-76.5)** |
| Female | **19.2 (18.1-20.3)** | **41.6 (40.3-42.9)** | **25.8 (23.5-28.2)** |
| **Race/Ethnicity** |  |  |  |
| Non-Hispanic White | **67.5 (66.1-68.8)** | 48.5 (47.3-49.7) | 49.1 (45.5-52.6) |
| Non-Hispanic Black | **15.8 (14.9-16.9)** | **26.2 (25.0-27.4)** | **32.1 (28.8-35.6)** |
| Non-Hispanic Asian | 2.0 (1.6-2.5) | 2.1 (1.7-2.6) | 1.2 (0.7-2.1) |
| Non-Hispanic Other* | 0.9 (0.8-1.1) | 1.2 (1.0-1.5) | 0.9 (0.6-1.5) |
| Non-Hispanic multiple races | **2.1 (1.7-2.5)** | 3.8 (3.5-4.1) | 3.9 (3.1-4.8) |
| Hispanic | 11.7 (10.8-12.7) | **18.2 (17.3-19.2)** | 12.8 (11.0-14.8) |
| **Sexual Identity** |  |  |  |
| Heterosexual | **93.7 (93.1-94.3)** | **86.0 (85.0-86.9)** | **88.9 (87.0-90.6)** |
| Lesbian or gay | 2.2 (1.9-2.6) | 3.9 (3.4-4.6) | 3.0 (2.0-4.6) |
| Bisexual | **4.1 (3.5-4.6)** | **10.1 (9.3-10.9)** | **8.1 (6.7-9.7)** |
| **Education** |  |  |  |
| Less than high school | 13.6 (12.5-14.7) | 14.4 (13.6-15.3) | 16.0 (13.7-18.7) |
| High school diploma or equivalent | **25.7 (24.6-26.9)** | 28.7 (27.7-29.8) | 31.5 (28.7-34.5) |
| Some college | 31.1 (29.9-32.4) | **36.7 (35.5-38.0)** | 35.7 (32.3-39.3) |
| College graduate | **27.4 (26.2-28.6)** | 10.4 (9.4-11.5) | 9.0 (7.4-10.8) |
| 12-17 year old | 2.2 (2.0-2.4) | 9.7 (9.3-10.2) | 7.8 (6.6-9.1) |
| **Poverty Level** |  |  |  |
| Living in poverty | **16.6 (15.6-17.7)** | 25.7 (24.6-26.9) | 26.0 (23.0-29.2) |
| Income up to 2x federal threshold | **18.9 (17.8-20.0)** | 26.2 (25.2-27.3) | 27.4 (24.8-30.1) |
| Income greater than 2x threshold | **64.5 (63.1-65.9)** | 48.0 (46.5-49.6) | 46.7 (43.2-50.2) |
| **Serious Psychological Distress** |  |  |  |
| No | **91.4 (90.6-92.2)** | 82.8 (81.5-84.1) | 80.8 (78.1-83.2) |
| Yes | **8.6 (7.8-9.4)** | 17.2 (15.9-18.5) | 19.2 (16.8-21.9) |
| **Days Used Cigars (Mean)** | **7.7 (7.5, 8.0)** | **11.2 (10.9, 11.5)** | **13.8 (13.2, 14.4)** |
| **Brand Used Most Often\*** |  |  |  |
| Backwoods | **3.7 (3.3-4.2)** | **12.6 (11.0-14.5)** | **6.3 (5.0-8.0)** |
| Black & Mild | **23.9 (22.7-25.2)** | **11.1 (9.3-13.2)** | **40.4 (37.8-43.1)** |
| Dutch Masters | **1.5 (1.2-1.8)** | **11.5 (9.5-13.7)** | **4.4 (3.3-5.9)** |
| Swisher Sweets | **11.4 (10.5-12.3)** | **32.4 (30.2-34.7)** | **17.9 (15.9-20.2)** |
| White Owl | **2.2 (1.8-2.6)** | **15.1 (13.2-17.2)** | **7.2 (5.7-8.9)** |
| Cohiba | **4.7 (4.0-5.6)** | **0.3 (0.1-0.8)** | **1.4 (0.9-2.4)** |
| Romeo y Julieta | **4.7 (4.0-5.5)** | **0.1 (0.1-0.3)** | **1.1 (0.7-1.8)** |
| Other brands in survey | **33.5 (32.0-35.1)** | 13.8 (11.6-16.3) | 16.0 (13.3-19.1) |
| Brand not specified in survey | **14.4 (13.6-15.3)** | 3.1 (2.2-4.3) | 5.2 (4.1-6.6) |
| **Past 30-Day Cigarette Use** |  |  |  |
| No | **48.9 (47.4-50.4)** | **37.3 (36.0-38.5)** | **25.6 (22.7-28.7)** |
| Yes | **51.1 (49.6-52.6)** | **62.7 (61.5-64.0)** | **74.4 (71.3-77.3)** |
| **Past 30-Day Cannabis Use** |  |  |  |
| No | 79.2 (77.7-80.6) | -- | -- |
| Yes | 20.8 (19.4-22.3) |  |  |

This table shows weighted percentages with 95% confidence intervals. Bold indicates statistically significant difference across cigar-blunt use category. Notes: CI = confidence interval. \*Non-Hispanic Other combines American Indian or Alaskan Native and Native Hawaiian or Pacific Islander categories due to small cell counts. \*Brand was only asked of those responding to the cigar items, and not those responding to exclusive blunt use; this captures 2,488 of 13,346 in the exclusive blunt category.

### Analyses

We estimated the weighted frequency and prevalence of each cigar-blunt use behavior, accounting for the complex survey design as specified in the NSDUH Statistical Inference Report for five years of pooled data. We conducted separate analyses by age: youth 12 to 17 years old, young adults 18 to 25 years, and adults 26 years and older. The mean number of days of cigar use were computed for each cigar-blunt use category for the total sample and each age group. We then identified the sociodemographic and substance use characteristics associated with categories of cigar-blunt use using Rao-Scott chi-square tests. We determined differences to be significant if CIs didn’t overlap. Stata version 18.0 was used for all analyses.

## RESULTS

Between 2015 and 2019, 48.6% (95% CI=47.6-49.6) of people 12 and older who reported past-30-day cigar use exclusively smoked cigars (i.e., no blunt use); 44.3% (95% CI=43.3-45.3) exclusively smoke blunts; and 7.2% (95% CI=6.8-7.6) smoked both cigars and blunts (Figure 1). The prevalence across cigar-blunt use categories differed by age group, with exclusive blunt use the most prevalent cigar use pattern among youth (72.5% [95% CI=70.7-74.3]) and young adults (62.4% [95% CI=61.4-63.5]), and exclusive cigar use most prevalent among adults 26+ (61.2% [59.8-62.5]). The frequency of use differed by cigar-blunt category, with those exclusively smoking cigars consuming significantly fewer days in the past month (7.7 [95% CI=7.5-8.0]), compared to 13.8 (95% CI=13.2-14.4) days for those smoking both blunts and cigars, and 17.5 (95% CI=16.8-18.2) days for those exclusively smoking blunts.

**Figure 1.**
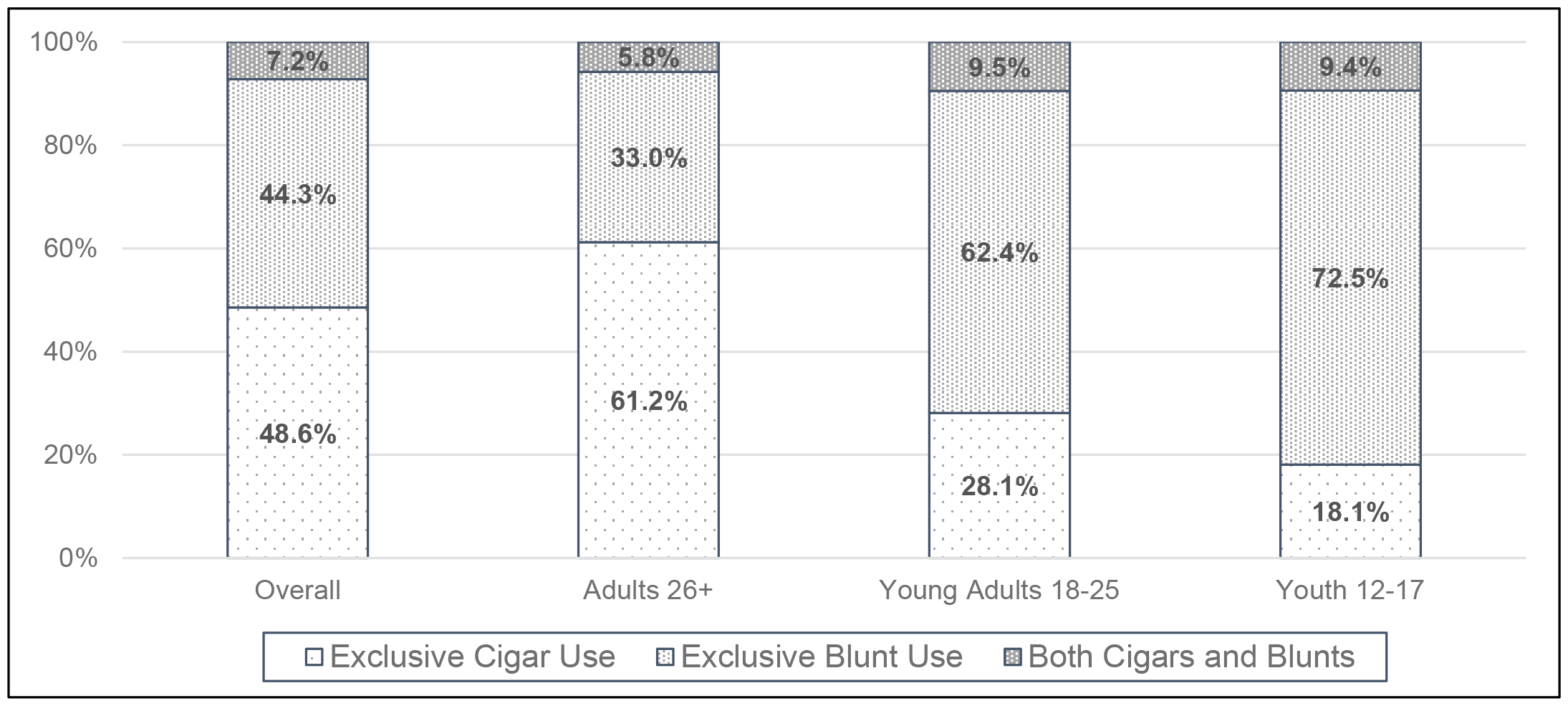
Cigar and blunt use patterns among youth and adults reporting past-30-day cigar use (N=25,004)

### Sociodemographic and Substance Use Characteristics Across Cigar-Blunt Use Category

There were significant differences in sociodemographic and substance use characteristics across the three cigar-blunt categories (Table 1). The proportion of youth (2.2%; 95% CI=2.0-2.4) and young adults (17.6%; 95% CI=6.6-9.1) was significantly lower within the exclusively cigar (no blunt) group compared to those exclusively smoking blunts (9.7% [95% CI=9.3-10.2] youth; 42.9% [95% CI=41.7-44.1] young adults) and both cigars and blunts (7.8% [95% CI=37.9-42.8] youth; 40.3% [95% CI=37.9-42.8] young adults). A greater proportion of exclusive blunt users (41.6%, 95% CI=40.3-42.9) reported being female compared to cigar and blunt users (25.8%, 95% CI=23.5-28.2) or exclusive cigar users (19.2%, 95% CI=18.1-20.3).

The racial/ethnic distributions differed significantly across groups, with a greater proportion of cigar and blunt users reporting being Black (32.1%; 95% CI=28.8-35.6) compared to exclusive blunt users (26.2%; 95% CI=25.0-27.4) and exclusive cigar users (15.8%; 95% CI=14.9-16.9); a greater proportion of exclusive blunt users reported Hispanic ethnicity (18.2; 95% CI=17.3-19.2) compared to cigar and blunt users (12.8; 95% CI=11.0-14.8) and exclusive cigar users (11.7; 95% CI=10.8-12.7). The proportion of participants reporting bisexual sexual identity was higher among those reporting exclusive blunt use (10.1%; 95% CI=9.3-10.9) compared to those reporting cigar and blunt use (8.1%; 95% CI=6.7-9.7) or exclusive cigar use (4.1%; 95% CI=3.5-4.6), with the opposite proportions identified for heterosexual identity. The proportion living in poverty was lower for those reporting exclusive cigar use (16.6%; 95% CI=15.6-17.7) compared to exclusive blunt use (25.7%; 95% CI=24.6-26.9) or cigar and blunt use (26.0%; 95% CI=23.0-29.2). Past month serious psychological distress was less prevalent among those reporting exclusive cigar use (8.6%; 95% CI=7.8-9.4) compared to exclusive blunt use (17.2; 95% CI=15.9-18.5) or cigar and blunt use (19.2%; 95% CI=16.8-21.9).

Cigar brand used most often differed across groups, with Swisher Sweets most prevalent among those reporting exclusive blunt use (32.4%; 95% CI=30.2-34.7), while Black & Milds were most prevalent among those reporting cigar and blunt use (40.4%; 95% CI=37.8-43.1) and exclusive cigar use (23.9%; 95% CI=22.7-25.2). Past 30-day cigarette use was highest among those reporting cigar and blunt use (74.4%; 95% CI=71.3-77.3), followed by those reporting exclusive blunt use (62.7%; 95% CI=61.5-64.0), and exclusive cigar use (51.1%; 95% CI=49.6-52.6).

## DISCUSSION

Among U.S. youth and adults who reported past 30-day cigar use, blunt use was prevalent, with over half of people reporting consuming cigars doing so exclusively or sometimes as blunts. Using cigars as blunts was particularly prevalent among youth and young adults, with nearly 3 in 4 youth and 3 in 5 young adults reporting exclusively using cigars as blunts. These findings align with other studies noting the prevalence of cannabis-tobacco co-use, particularly among youth.^4^

In the U.S., cigars are co-marketed with cannabis descriptors,^21^ with industry-targeted marketing more prevalent in neighborhoods aligning with the sociodemographic factors identified in this study as more common among those reporting blunt use (e.g., by race, socioeconomic status, sexual orientation). The identified differences in sociodemographic and substance use characteristics by cigar-blunt use groups suggest that youth and adults who use cigars as blunts, whether exclusively or sometimes, are more similar to each other than those who do not use cigars as blunts. Additional factors such as higher prevalence of cigarette use and more days of cigar use per month indicate that these two groups may be at greater risk for negative health outcomes related to combusted tobacco use. This latter finding aligns with other studies that found that adults who use both cigars and blunts have more severe risk profiles.^19^

There are several public health implications for these findings. First, studies that aggregate cigars and blunts into one group may limit potentially meaningful subgroup risk profiles. Second, when assessing cigar use, particularly among youth and young adults, it is important to consider blunt use. With youth not considering blunts as cigar products,^9,10,19^ definitions may be needed for accurate measurement. Additionally, including blunts in cigar measurement may allow for more accurate tobacco regulation. Blunts are typically purchased as cigar products, with the modifications taking place after purchase by the end user. As such, the manufacturing, sale, and distribution of blunts in the U.S. would be regulated through the U.S. Food and Drug Administration as a cigar product. Policies that regulate cigars are likely to influence blunt use, though, without accurate measurement of cigar use, including blunt use, these impacts may be overlooked. Finally, given the high rates of blunt use among youth and young adults identified in the present study, along with findings from other studies indicating increased likelihood for continued and co-use with other tobacco products,^15^ additional education efforts may be warranted for this population to reduce long-term risks.

Study limitations should be considered when interpreting findings. First, NSDUH is representative of the U.S. civilian population, and findings may not represent non-civilian and institutionalized people. Second, due to the cross-sectional nature of the study, we cannot make claims of causality. Third, data are self-reported and subject to response bias. Indeed, there were inconsistencies identified among those reporting the number of days consuming blunts and cigars, particularly among those reporting exclusively using cigars as blunts. However, a conservative approach was taken in rectifying inconsistencies. Fourth, other tobacco products, including e-cigarettes and hookah, were not assessed during the study period, which limits our ability to further explore patterns of dual and poly use. Finally, the analyses were limited to those reporting past 30-day cigar use based on the available measure for replacing cigars with cannabis. Cigar use, particularly premium cigar use, is noted to be less frequently reported.^22^ The identified patterns may not reflect less-frequent cigar-blunt use.

Blunt use is common among U.S. youth and adults who smoke cigars. Indeed, the majority of youth aged 12-17 and young adults 18-24 who use cigars report exclusively using cigars as blunts. Further, youth and adults who use cigars as blunts reported smoking more cigars per month, increasing their risk for negative health outcomes. Sociodemographic and substance use characteristics suggest those using cigars exclusively as blunts or sometimes as blunts are more similar than those not reporting any blunt use. These findings have important implications for measurement and education.

## Data Availability

We analyzed publicly available NSDUH data. Analytic code available upon reasonable request to the authors.

https://www.samhsa.gov/data/data-we-collect/nsduh-national-survey-drug-use-and-health

---

**Supplemental Table 1.** NSDUH Variables Used in Analyses.

| **Supplemental Table 1. NSDUH Variables Used in Analyses** |  |  |
| --- | --- | --- |
| **Construct** | **Variables Used** | **Recoding Notes** |
| Past 30-day cigar/blunt use | CIGARREC<br>BLNTREC<br>BLNTNOMJ | Used to select sample who reported past 30-day cigar or blunt use |
| Cigar-cannabis use | CIGARREC<br>BLNTREC<br>BLNTNOMJ<br>MJDAY30A<br>BLNT30C1<br>BLNT30C2 | Exclusively cigars – none of the cigars smoked in the past 30 days had cannabis<br>Exclusively blunts – all cigars smoked in the past 30 days had cannabis<br>Both cigars and blunts – smoked at least one cigar with cannabis in the past 30 days, but not all<br>BLNT30C1 and BLNT30C2 were used to resolve conflicting responses when reported blunt and cannabis use disagreed. |
| Age | CATAG6 | None |
| Sex | IRSEX | None |
| Race/Ethnicity | NEWRACE2 | Combined American Indian or Alaskan Native and Native Hawaiian or Pacific Islander categories due to small cell counts |
| Sexual identity | SEXIDENT | None |
| Education | EDUHIGHCAT | None |
| Poverty level | POVERTY3 | None |
| Serious psychological distress | SPDMON | None |
| Days used | CGR30USE<br>BLNT30DY | For those with responses to both items, the highest was used to estimate mean days used by group |
| Cigar brand used most often | CGR30BR2 | Less frequently reported brands were combined |
| Past 30-day cigarette use | CIGREC | Recoded those that reported use within the past 30 days as 1 and all others as 0. |
| Past 30-day cannabis use | MJREC | Recoded those that reported use within the past 30 days as 1 and all others as 0. |
| Note: Analytic code available upon reasonable request to corresponding author. |  |  |

## Notes

Funding: Research reported in this publication was supported by grant numbers K01CA253235 (JKJ), U01CA278695 (CDD), and U54CA229973 (CDD), from the National Cancer Institute and the FDA Center for Tobacco Products. The content is solely the responsibility of the authors and does not necessarily represent the official views of the NIH or the FDA.

### Competing Interest Statement

The authors have declared no competing interest.

### Funding Statement

Research reported in this publication was supported by grant numbers K01CA253235 (JKJ), U01CA278695 (CDD), and U54CA229973 (CDD), from the National Cancer Institute and the FDA Center for Tobacco Products. The content is solely the responsibility of the authors and does not necessarily represent the official views of the NIH or the FDA.

